# Quantification of the association between predisposing health conditions, demographic, and behavioural factors with hospitalisation, intensive care unit admission, and death from COVID-19: a systematic review and meta-analysis

**DOI:** 10.1101/2020.07.30.20165050

**Authors:** Nathalie Verónica Fernandez Villalobos, Jördis Jennifer Ott, Carolina Judith Klett-Tammen, Annabelle Bockey, Patrizio Vanella, Gérard Krause, Berit Lange

## Abstract

**Background:** Comprehensive evidence synthesis on the associations between comorbidities and behavioural factors with hospitalisation, Intensive Care Unit (ICU) admission, and death due to COVID-19 is lacking leading to inconsistent national and international recommendations on who should be targeted for non-pharmaceutical interventions and vaccination strategies.

**Methods:** We performed a systematic review and meta-analysis on studies and publicly available data to quantify the association between predisposing health conditions, demographics, and behavioural factors with hospitalisation, ICU admission, and death from COVID-19. We provided ranges of reported and calculated effect estimates and pooled relative risks derived from a meta-analysis and meta-regression.

**Results:** 75 studies were included into qualitative and 74 into quantitative synthesis, with study populations ranging from 19 - 44,672 COVID-19 cases. The risk of dying from COVID-19 was significantly associated with cerebrovascular [pooled RR 2.7 (95% CI 1.7-4.1)] and cardiovascular [RR 3.2 (CI 2.3-4.5)] diseases, hypertension [RR 2.6 (CI 2.0-3.4)], and renal disease [RR 2.5 (CI 1.8-3.4)]. Health care workers had lower risk for death and severe outcomes of disease (RR 0.1 (CI 0.1-0.3). Our meta-regression showed a decrease of the effect of some comorbidities on severity of disease with higher median age of study populations. Associations between comorbidities and hospitalisation and ICU admission were less strong than for death.

**Conclusions:** We obtained robust estimates on the magnitude of risk for COVID-19 hospitalisation, ICU admission, and death associated with comorbidities, demographic, and behavioural risk factors. We identified and confirmed population groups that are vulnerable and that require targeted prevention approaches.

**Summary:** Comorbidities such as cardiovascular disease or hypertension are less strongly associated with hospitalization and ICU admission than with death in COVID-19 patients. Increasing age is associated with a lower effect on comorbidities on disease severity.

## 1. Introduction

Various factors have been identified to determine whether a higher risk of severe course of COVID-19 disease and COVID-related deaths exists. Some of these identified factors are demographic in nature, such as age and sex, others have to do with having a diagnosed condition such as diabetes and hypertension (1–5). Furthermore, behavioural and occupational risk factors have also been discussed (6–9). Accordingly, during the pandemic, part of non-pharmaceutical interventions were aimed towards these corresponding population groups. For example, several affected countries worldwide have recommended to avoid visiting elderly relatives for recreational reasons (10).

Public health officials as well as national guidelines have been inconsistent and sometimes vague in defining exact target groups for health measures other than the elderly (11,12). Part of the reason for this is that many studies have reported some of these predisposing factors using incomparable data sources, indicators and calculations, and denominators. Estimates are further challenged by the interrelation between factors such as age and comorbidities, which limits causal and relative attributions (13–15). Several studies published have refrained from giving effect estimates for factors experienced by patients but have only described these data in the form of clinical case series (16–18).

To inform national and international guidelines and policies on the actual impact of targeting different population groups with predisposing conditions or factors evidence synthesis is needed to quantify the risk of individual predisposing factors and conditions. This will inform both toward protection of potential risk groups, and on potential vaccination strategies in the case of not-optimal availability or efficacy of potential vaccination candidates.

However, currently there is only fragmented evidence synthesis available on quantifiable information on the actual risk experienced by patients with these comorbidities for the most important outcomes. Existing systematic reviews on that subject have been published either as pre-print or as peer reviewed publications to date (6–8,19–23). Some of these focus on one association to a particular endpoint, e.g. cardiovascular morbidity and severity of course of disease (24–26). Some assess several comorbidities as well (4,23), but do not cover many studies yet. Moreover, these do not investigate the interaction of age or do not account for those studies that do not report associations (this information is available in the Supplement, table 1). To our knowledge, no existing evidence synthesis provides comparative and exhaustive relative risk measures of the major predisposing conditions in most populations, using both data with and without reported relative risk measures, taking age structure of the respective study population into account.

We conducted a meta-analysis on risk for COVID-related hospitalisation, Intensive Care Unit (ICU) admission, and death and its association with comorbidities, behavioural factors, and death. We extracted and included crude study data to understand the magnitude of the effect of comorbidities and other factors on COVID-19 health outcomes. By focussing on these outcomes, our objective was to generate evidence for prioritising health measures for vulnerable population groups. Our results shall also establish baseline information to identify priority groups for COVID-19 intervention strategies, e.g. vaccination.

## 2. Material and methods

### Search strategy

We performed a systematic review (registration number in PROSPERO CRD42020190548), following PRISMA guidelines (27), in MEDLINE, bioRXiv, and MedRXiv, searching for publications on COVID-19 and risk groups for severe or lethal disease outcomes (search terms “novel coronavirus”, “COVID-19”, “SARS-CoV-2”). We applied the snowball method to available systematic reviews to identify further evidence. The existing reviews are listed in the Supplement, table 1. The literature search includes reports up to 29 April 2020.

In addition to that search, we identified reports from other publicly available sources, namely national (public) health institutions, and data repositories, with a last search date of 28 May 2020.

### Inclusion and exclusion criteria

We included reports if: a) patients had COVID-19, either confirmed microbiologically or clinically (population); b) information on COVID-19 outcome was reported as either death (hospital or after a defined follow-up time), ICU admission (both ICU and intermediate care), hospitalisation or aggravation of disease (clinical description) (outcome); and c) at least one comorbidity, risk factor or behavioural factor was described and if number of patients with/without outcome was reported according to the respective factor (exposure and comparison).

Eligible study designs were: cohort studies, cross-sectional studies, case series, and clinical trials. Languages included were English, Spanish, Italian, French, or German.

The data included from country-level reports focus on the seven countries with the highest officially reported absolute number of deaths due to COVID-19 by 28 April 2020^1^, according to numbers of the Johns Hopkins University (28). We used data on age, sex, and comorbidities of reported cases. The outcomes were hospitalisations and death.

### Data extraction

We developed spreadsheets for crude data extraction. The template was developed and tested by the three extracting scientists (CK, NF, JO).

We extracted relevant variables in the smallest reported unit and according to the main stratification variable, either comorbidity or behavioural risk factor, author and link, country, data source, age-range, study time-frame, baseline population group, outcome (mortality, severity, or other), number of individuals in the risk group, total sample, number of individuals among risk group with outcome, total number of individuals people with the outcome, and effect measures of association reported as well as relative risks computed automatically.

The outcomes were severity/aggravation during the course of COVID-19 disease in terms of hospitalisation/pneumonia, admission to ICU, and death. “Risk groups” were those with comorbidities, which we defined and grouped according to the International Classification of Diseases 11^th^ Revision (ICD-11) (https://icd.who.int/browse11/l-m/en). We extracted the five most common comorbidities or behavioural/occupational/demographic factors per included study.

For data from research reports we did random plausibility checks and plotted relative risks with ranges. A researcher not involved in data extraction (AB) double-checked 20% of included studies and compared extracted numbers with original reports.

For publicly available data we extracted data for all seven countries on: age of confirmed COVID-19 cases, hospitalisations, ICU admissions and deaths. For the US, Spain, and France we additionally extracted mortality data, distinguished by comorbidities, which we used to estimate relative risk of death for cases, or in the case of France, for hospitalisations.

### Risk of bias

We assessed risk of bias using an adapted version of the ROBINS-I tool (29) for non-randomised studies. We analysed the studies in terms of bias due to confounding, selection of participants and follow-up, misclassification of exposure, missing data, measurement of outcome, or reporting. We measured the risk scales as low, moderate, and high.

### Data analysis

#### Descriptive

We display ranges of reported estimates of association [Odds ratios (ORs), Hazard ratios (HRs), and Relative Risks (RRs)] for the health outcomes from included studies and calculate relative risks (RRs) for each risk group and for each outcome, based on crude and absolute data from the studies. Strata of outcomes that reported “zero” were excluded from the analysis, as this gives an invalid statistical estimate of the underlying risks.

For data from publicly available sources, we computed point estimates of the relative risks to severe health status, like hospitalisation or death; if possible, stratified by age groups and sex with 95% confidence intervals (CIs). In addition, we estimated relative risks of death among three age groups and two sexes for cases in Spain and hospitalised cases in France.

#### Meta-analysis and meta-regression

We assessed heterogeneity visually in forest plots and by assessing the percentage of variance over studies I^2^. Due to the difference between populations and observed heterogeneity, we performed a random effects meta-analysis for pooled RRs. For those risk groups with considerable heterogeneity (>75%), we performed subgroup analyses to investigate reasons for heterogeneity further according to the Cochrane Handbook for Systematic Reviews of Interventions (https://training.cochrane.org/handbook/current). Within meta-regression, we assessed effect modification by age on relative risks of included comorbidities or other risk factors.

In order to include explanatory variables, we ran a series of mixed-effects meta-regressions to test the associations of case mortality of COVID-19 with demographics and different comorbidities. We included 11 of the studies presented in this paper (18,30–39) that reported data on the variables of interest. Our baseline modelling approach (40) for this was:

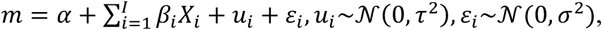

with *X*_*i*_ being the *i*^*th*^ moderator variable, *τ* being the homogeneity estimate among the studies and *ε*_*i*_ being an independent white noise process. The majority of studied individuals had comorbidities like cardiovascular disease, diabetes, or hypertension.

We checked the correlation matrix among the variables median age, share of males, share of individuals with cardiovascular disease, share of individuals with diabetes, and share of individuals with hypertension.

As correlations between cardiovascular disease and hypertension with diabetes were especially high, diabetes was ignored to avoid high multicollinearity among the regressors. To take collinearity between age and the comorbidities into account, we included interactions between the median age, the share of individuals with cardiovascular disease, and the share of individuals with hypertension in our full model.

We iteratively omitted variables with lowest p-values and compared the models using the log-likelihoods, the corrected version of Akaike’s Information Criterion (AICc) and the Bayesian Information Criterion (BIC). Model fit was performed in **R** using the **rma**.**uni()** command in the **metafor** package (40).

## 3. Results

We identified a total of 7,429 records. We retrieved 190 of them for full-text screening and 75 studies met the inclusion criteria (Supplement, figure 1). All of them were used for our qualitative analysis, and 74 for the quantitative analysis. The pre-print study “OpenSAFELY: factors associated with COVID-19-related hospital death in the linked electronic health records of 17 million adult NHS patients” by Williamson et al. was excluded because the actual number of confirmed COVID-19 cases was not reported.

The majority of reports (n=66) were from China, followed by the United States of America (USA, n=5). Studies were based on medical or clinical records (n=69) or official reported data (n=4) and conducted between late December 2019 and April 2020 with follow-up of 5-30 days. Endpoints were: hospitalisation/pneumonia (n= 43), admission to ICU (n=17), and death (n=26; mostly within 30 days or in-hospital deaths). Three studies had composite endpoints, three others reported multiple endpoints.

The sample sizes were between 19 and 44,672 confirmed COVID-19 cases and individuals were aged between 33–82 years; and in seven studies children were included (supplement, table 2).

### 3.1. Risk of bias assessment

We assessed risk of bias due to a) confounding, b) selection, c) misclassification, d) missing data, and e) measurement of outcome. Confounding was moderate in most studies as either adjusted estimates or age-information was provided. Selection bias was mostly low to moderate as was misclassification. A high risk of bias was found for 45 studies due to non-reporting or missing data. In several studies the source of data or definition of the outcome was unclear, and several reported results in selective subgroups. In general, studies had at least one moderate risk in terms of follow-up or adjusting for confounders and only few had overall low risk of bias in all categories (supplement, table 3).

### 3.2. Reported and calculated associations

Fourteen studies reported odds ratios, nine studies reported hazards ratios, and one study reported relative risks for either one of the endpoints included. One study included cases reported from official data. We calculated RRs from crude study data for a) hospitalisation/pneumonia, b) admission to ICU, and c) death (within 30 days or within hospital) for all 74 studies included in the quantitative analysis (Table 1).

**Table 1.**
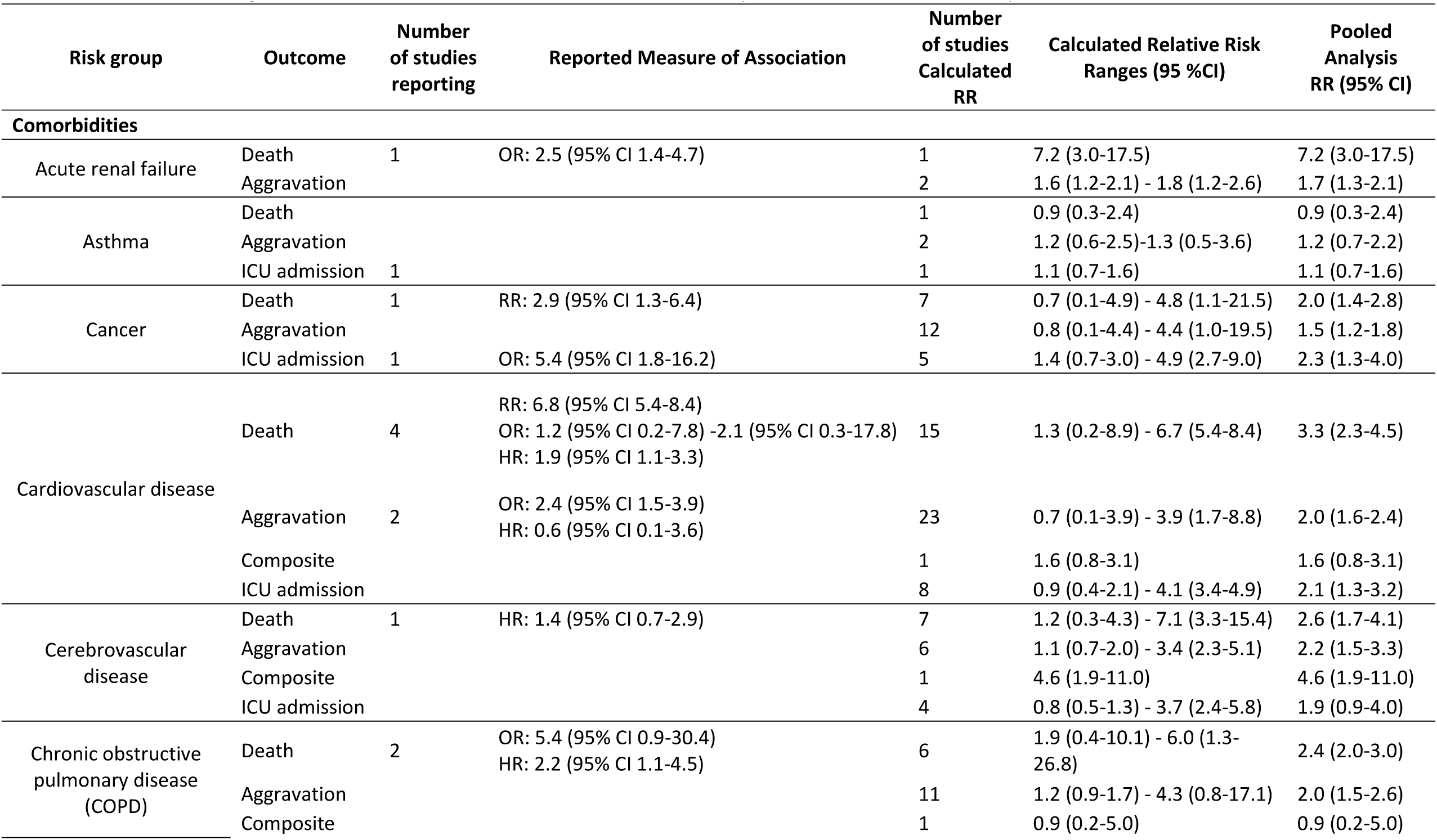
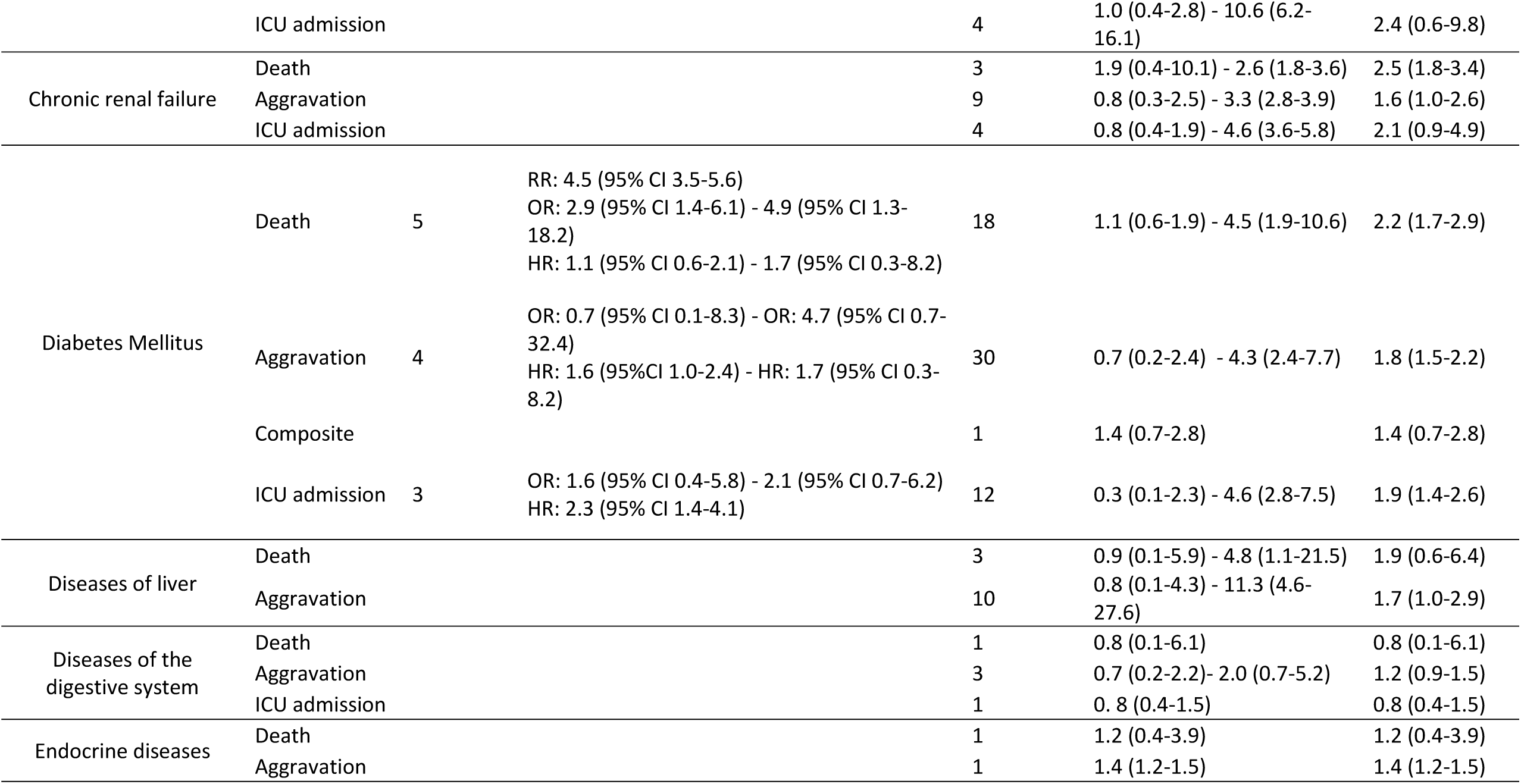
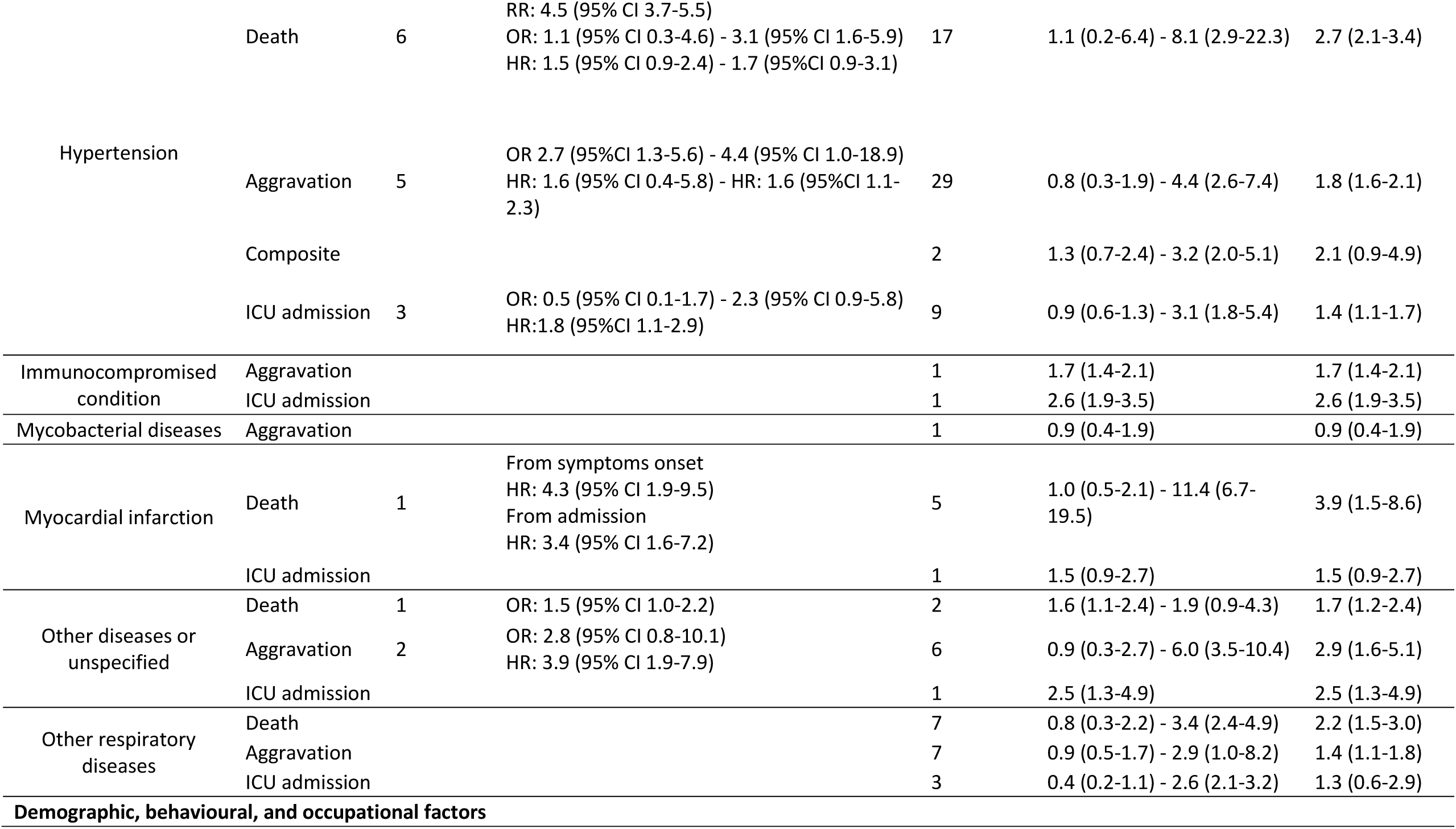
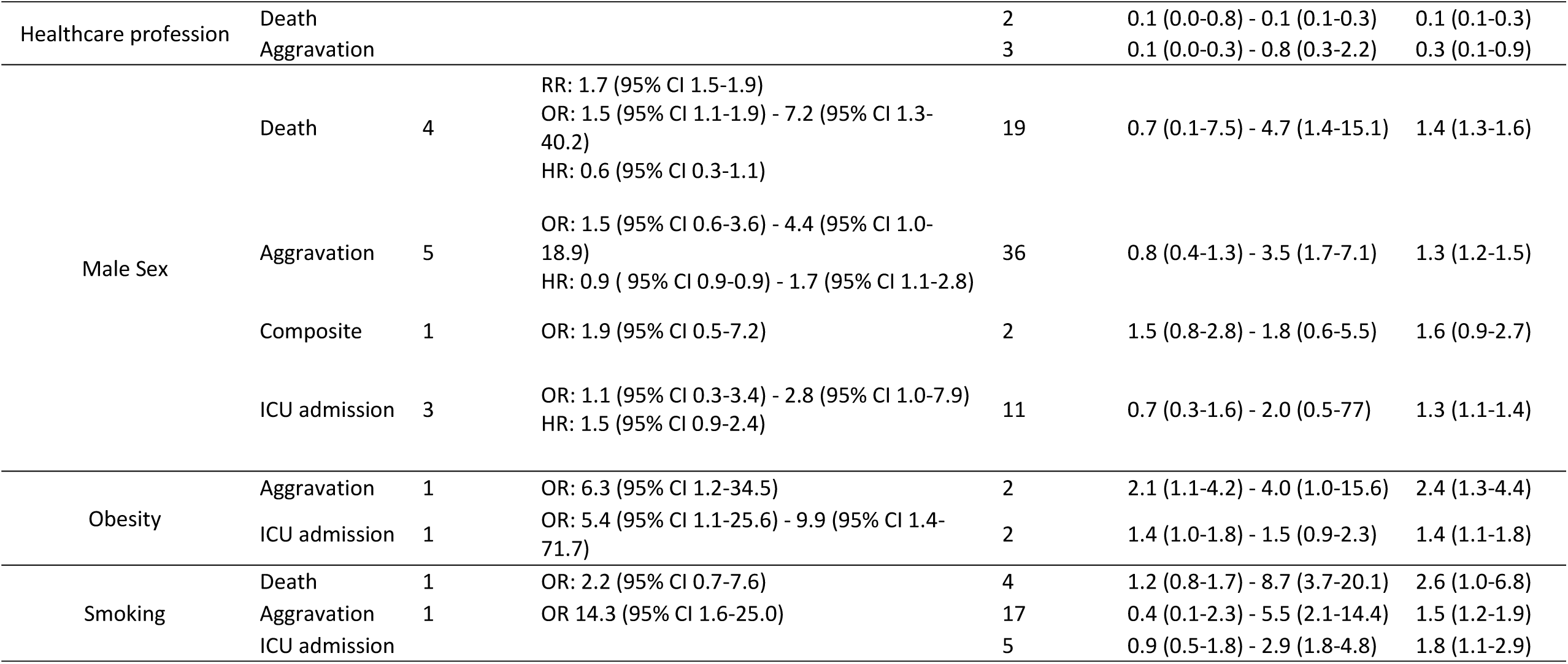
Association in original studies and own calculations of relative risks by COVID-19 outcomes and by risk

#### 3.2.1. Hospitalisation

Three studies reported higher odds of being hospitalised due to COVID-19 for patients with acute renal failure, cardiovascular disease, and diabetes [Odds Ratio 2.5 (95% CI 1.4-4.7); Odds Ratio 2.4 (CI 1.5-3.9); Hazard Ratio 1.6 (CI 1.0-2.4), respectively]. Hypertension was reported as a factor that increased the odds of being hospitalised [Odds Ratio 2.7 (CI 1.3-5.6) - 4.4 (CI 1.0-18.9), Hazard Ratio 1.6 (CI 1.1-2.3), 5 studies] (Table 1).

Using crude data from all clinical case series that provided these numbers, we calculated that patients with acute renal failure, cerebrovascular disease, and COPD had higher risk of hospitalisation [RR 1.6 (95% CI 1.2-2.1) - 1.8 (CI 1.2-2.6), 2 studies], [1.1 (CI 0.7-2.0) - 3.4 (CI 2.3-5.1), 6 studies [1.2 (CI 0.9-1.7) - 4.3 (CI 0.8-17.1), 11 studies], respectively (Table 1).

Regarding the association of demographic, behavioural, and occupational factors and hospitalisation, four studies reported that male patients had higher odds and hazard of being hospitalised [OR 3.7 (95%CI 1.75-7.75), HR 1.7 (CI 1.0-2.8)], while two studies did not see evidence of association [OR 0.5 (CI 0.3-0.8), HR 0.9 (CI 0.9-0.9)] (Table 1).

Using the crude data, we found that male patients [RR 1.1 (95% CI 1.0-1.2) - 3.5 (CI 1.7-7.1) 29 studies], patients who smoke [1.1 (CI 0.4-2.8) - 5.5 (CI 2.1-14.4), 14 studies], and obese patients [2.1 (CI 1.1-4.2) - 4.0 (CI 1.0-15.6), 2 studies] had higher risk of disease aggravation. Other studies did not find associations between being male [0.8 (CI 0.4-1.3) - 0.9 (0.6-1.6), 7 studies] or smoking [0.4 (CI 0.1-2.3) - 0.7 (CI 0.2-1.8), 3 studies] and hospitalisation. Healthcare workers were found to have a lower risk of being hospitalised due to COVID-19 [RR 0.1 (CI 0.0-0.3) - 0.8 (CI 0.3-2.2), 3 studies] (Table 1).

#### 3.2.2. ICU admission

Four studies reported on those patients that needed to be admitted at ICU based on medical records. Three studies reported cancer, diabetes, and hypertension as factors that increase the odds of being admitted at ICU [OR 5.4 (95% CI 1.8-16.2), HR 2.3 (CI 1.4-4.1), HR 1.8 (CI 1.1-2.9), respectively] (Table 1).

Using crude numbers of patients and events provided in studies, we calculated that patients with cancer [RR 1.4 (95% CI 0.7-3.0) - 4.9 (CI 2.7-9.0), 5 studies] and COPD [1.0 (CI 0.4-2.8) - 10.6 (CI 6.2-16.1), 4 studies] had a high risk of being admitted at ICU (Table 1).

Regarding other risks, one study reported that male patients [OR 2.8 (95 %CI 1.0-7.9)] had higher odds of being admitted at ICU or having invasive mechanical ventilation (Table 1).

We found obesity [RR 1.4 (95% CI 1.0-1.8) - 1.5 (CI 0.9-2.3), 2 studies], male gender [RR 1.1 (CI 0.7-1.8)-2.0 (CI 0.5-7.7), 9 studies], and smoking [RR of 2.4 (CI 1.1-2.9) - 2.9 (CI 1.8-4.8), 3 studies] to be a risk factor for ICU admission. Other studies did not find associations between being male [0.7 (CI 0.3-1.6) - 0.9 (CI 0.6-1.4), 2 studies] or smoking [0.9 (CI 0.5-1.8) - 0.9 (CI 0.6-1.5), 2 studies] and ICU admission (Table 1).

#### 3.2.3. Death

Eight studies reported on patients deceased due to COVID-19, and one study included cases reported from official data (Table 1). Four studies reported acute renal failure, cancer, COPD, and myocardial infarction as comorbidities associated with death [OR 2.5 (95% CI 1.4-4.7), RR 2.9 (CI 1.3-6.4); HR: 2.2 (CI 1.1-4.5), and HR 3.4 (CI 1.6-7.2), respectively]. Patients with cardiovascular disease [HR 1.9 (CI 1.1-3.3), RR 6.8 (CI 5.4-8.4), 4 studies], diabetes [OR 2.9 (CI 1.4-6.1) - 4.9 (CI 1.3-18.2), RR 4.5 (CI 3.5-5.6), 5 studies], and hypertension [OR 3.1 (CI 1.6-5.9), HR of 1.6 (CI 1.1-2.3), RR of 4.5 (CI 3.7-5.5), 6 studies] had higher risk of dying because of COVID-19 (Table 1).

Calculating from crude numbers of patients and events provided in study reports, we found that patients with cardiovascular disease [RR 1.3 (95% CI 0.2-8.9) - 6.7 (CI 5.4-8.4), 15 studies], cerebrovascular disease [1.2 (CI 0.3-4.3) - 7.1 (CI 3.3-15.4), 7 studies], COPD [1.9 (CI 0.4-10.1)-6.0 (CI 1.3-26.8), 6 studies], diabetes [1.1 (CI 0.6-1.9)- 4.5 (CI 1.9-10.6), 18 studies], and hypertension [1.1 (CI 0.2-6.4)-8.1 (CI 2.9-22.3), 17 studies] had a higher risk of dying due to COVID-19 than those patients without these comorbidities (Table 1).

Regarding demographic, behavioural, and occupational factors, two studies reported that male patients had higher odds of death [OR 1.5 (95% CI 1.1-1.9), RR 1.7 (CI 1.7-1.9)] (Table 1).

Based on the calculated associations, we found that male patients [RR 1.2 (95% CI 0.9-1.6) - 4.7 (CI 1.4-15.1), 14 studies] and patients who smoke [RR 1.2 (CI 0.8-1.7) - 8.7 (CI 3.7-20.1), 4 studies] had higher risk of dying due to COVID-19. However, based on five studies, we did not find any association between being male and death [0.7 CI (0.1-7.5) - 0.9 (CI 0.6-1.4)]. Healthcare workers were found to have less risk of death [RR 0.1 (CI 0.0-0.8) - 0.1 (CI 0.1-0.3), 2 studies] (Table 1).

##### National primary care electronic health record data linked to in-hospital COVID-19 death data

The study conducted by Williamson et al. (OpenSAFELY: factors associated with COVID-19-related hospital death in the linked electronic health records of 17 million adult NHS patients) reported their results based on primary care records for patients in England. They found that death from COVID-19 was associated with being male [HR 1.9 (95%CI 1.8-2.1)], age [HR 2.1 (1.8-2.4), HR 4.8 (4.2-5.4), and HR 12.6 (11.2-14.3) for age groups 60-69, 70-79, >80 years of age, respectively taking into account age group 50-59 as a reference], uncontrolled diabetes [HR 2.3 (CI 2.1-2.5)], and severe asthma [HR 1.2 (CI 1.0-1.4)]. This study was not included in our meta-analysis as it did not provide absolute numbers of patients infected with COVID-19.

### 3.3. Meta-analysis and meta-regression: Association of comorbidities and demographic, behavioural, and occupational factors with hospitalisation, ICU admission and death; effect modification of age

We performed random-effects meta-analysis on the influence of comorbidities and other factors on three endpoints (hospitalisation, ICU admission, and death) (figure 1). To assess the influence of age on these associations we performed meta-regressions for those associations with more than 15 studies available on the influence of age on these associations. We also performed a mixed-effects meta-regression on the main comorbidities adjusted for median/mean age and gender.

**Figure 1.**
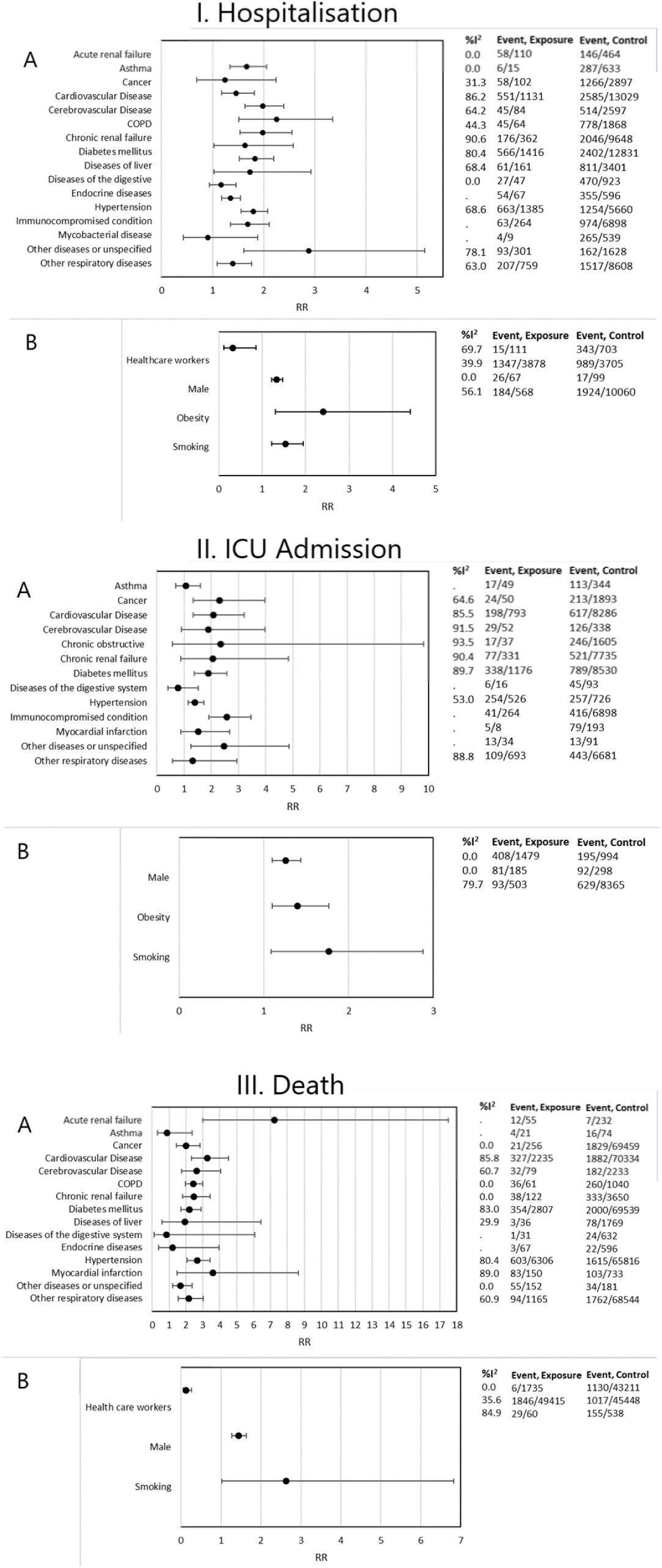
Summary effect meta-analysis of hospitalisation, ICU admission, and death. **I. Hospitalisation:** A. association of hospitalisation and comorbidity, B. association of hospitalisation and demographic, behavioural, and occupational. **II. ICU admission:** A. association of ICU admission and comorbidity, B. association of ICU admission and behavioural, demographic and occupational factors. **III. Death:** A. association of death and comorbidity, B. association of death and behavioural, demographic and occupational factors.

#### 3.3.1. Hospitalisation

##### Comorbidities

The random-effects meta-analysis found patients with cardiovascular disease (RR 1.9, 95% CI 1.6-2.4), cerebrovascular disease (RR 2.3, 95% CI 1.5-3.3), and/or diabetes mellitus (RR 1.8, 1.5-2.6) were at higher risk of hospitalisation. Other comorbidities, including chronic renal disease, chronic respiratory disease, and COPD, were correlated with higher hospitalisation rates as well. We found moderate to high heterogeneity for several of these risk factors (I^2^ 50-90%) (Figure 1.I.A.).

##### Demographic, behavioural, and occupational factors

In pooled results from random effect meta-analysis obese individuals had 2.4 times the risk of being hospitalised compared to those without obesity (RR 2.4, 95% CI 1.3-4.4). Healthcare workers were less likely to be hospitalised (RR 0.3, 95% CI 0.1-0.9), and males were 30% more likely to be hospitalised than females (RR 1.3, 95% CI 1.2-1.5) (Figure 1.I.B.).

#### 3.3.2. ICU admission

##### Comorbidities

The following comorbidities were associated with high risk for ICU admission (Figure 1.II.A.) in pooled analysis: cancer (RR 2.3, 95%CI 1.3-4), diabetes mellitus (RR 1.9, CI 1.4-2.6), and cardiovascular conditions (RR 2.0 95%CI 1.3-3.2). Heterogeneity of pooled results was moderate to high.

##### Demographic, behavioural, and occupational factors

Obesity and smoking moderately increased the risk to being admitted to ICU (Figure 1.II.B.). Information on healthcare workers was insufficient to pool results.

#### 3.3.3. Death

##### Comorbidities

The highest observed RR of death were found for cerebrovascular disease, cardiovascular disease, chronic renal disease, and hypertension (RR 2.7 95%CI 1.7-4.0), (3.2, CI 2.3-4.5), (2.5, CI 1.8-3.4), and (2.6, CI 2.0-3.4), respectively (Figure 1.III.A.).

##### Demographic, behavioural, and occupational factors

Males had risk of death due to COVID-19 1.4 times that of females (95% CI 1.3-1.6), and healthcare professionals were at lower risk of death due to COVID-19, when compared to other population groups (RR 0.12, CI 0.06-0.27) (Figure 1.III.B).

#### 3.3.4. Effect modification

##### Hospitalisation

The meta-regression revealed that the strength of the association between comorbidities and hospitalisation decreased with increased median or mean age of the study population [cardiovascular (beta-Coefficient −0.05, 95%CI – 0.09-0.003, p=0.038) and diabetes (beta-Coefficient −0.07, 95%CI −0.1- - 0.03, p=0.002)], (Figure 2.A).

**Figure 2.**
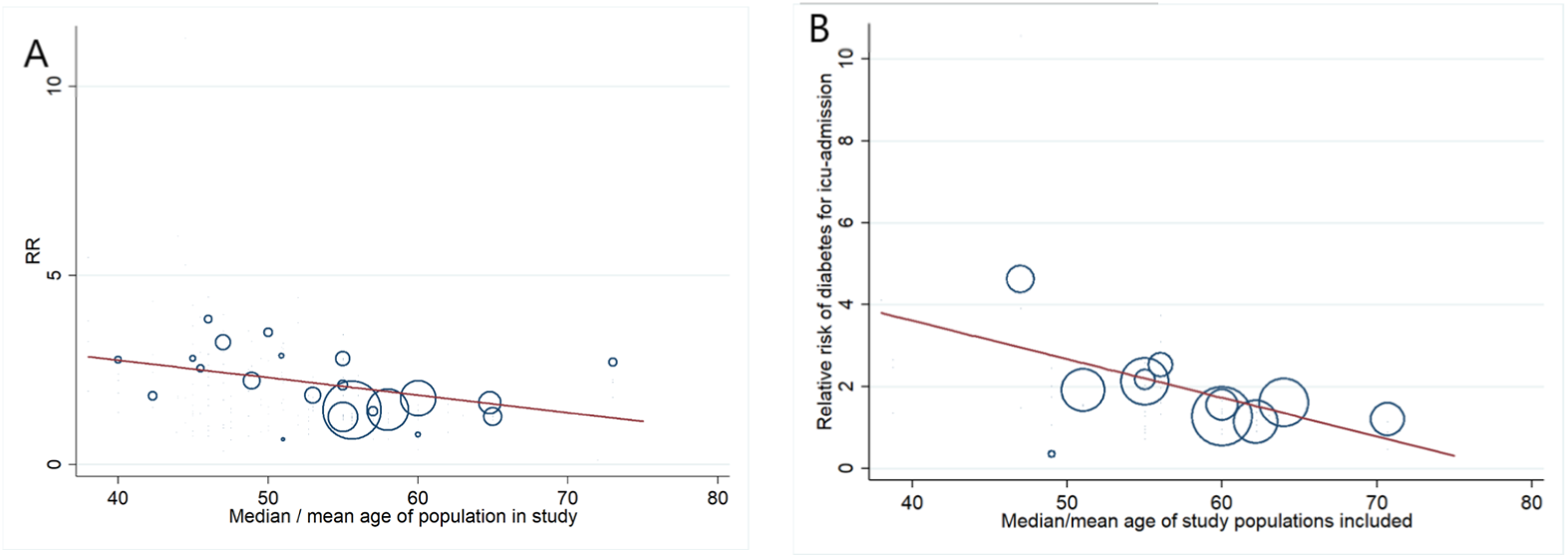
Effect modification of mean/median age of population in study: A. modification of association between cardiovascular morbidity and hospitalisation, B. modification of age on the association of diabetes and ICU admission risk.

##### ICU

Age was also modifying the association with diabetes; here, the RRs for ICU admission decreased with increasing age (beta-coefficient −0.1, 95%CI −0.2- - 0.004, p=0.042) (Figure 2.B). We did not find effect modification of age for other risk factors like gender, hypertension, or smoking.

##### Death

Effect modification was found for the association of hypertension with dying from COVID-19 with higher relative risks in those studies with lower median/mean ages (beta-coefficient - 0.14, 95%CI −0.27- - 0.022, p= 0.025). We did not find effect modification for diabetes or cardiovascular morbidity.

#### 3.3.5. Meta-regression

In a meta-regression adjusted for gender and age (Table 2), results of Model 4 show the effect of age and comorbidities on mortality. The increase in mean mortality risk is more pronounced in older populations. For example, for a population with a median age of 50 years, a 1%-increase in prevalence of hypertension is associated with an increase in the overall mortality rate 0.7%^2^. For hypertension, this increase would result in a mortality rate of 0.9% for a median age of 70 years^3^.

**Table 2.**
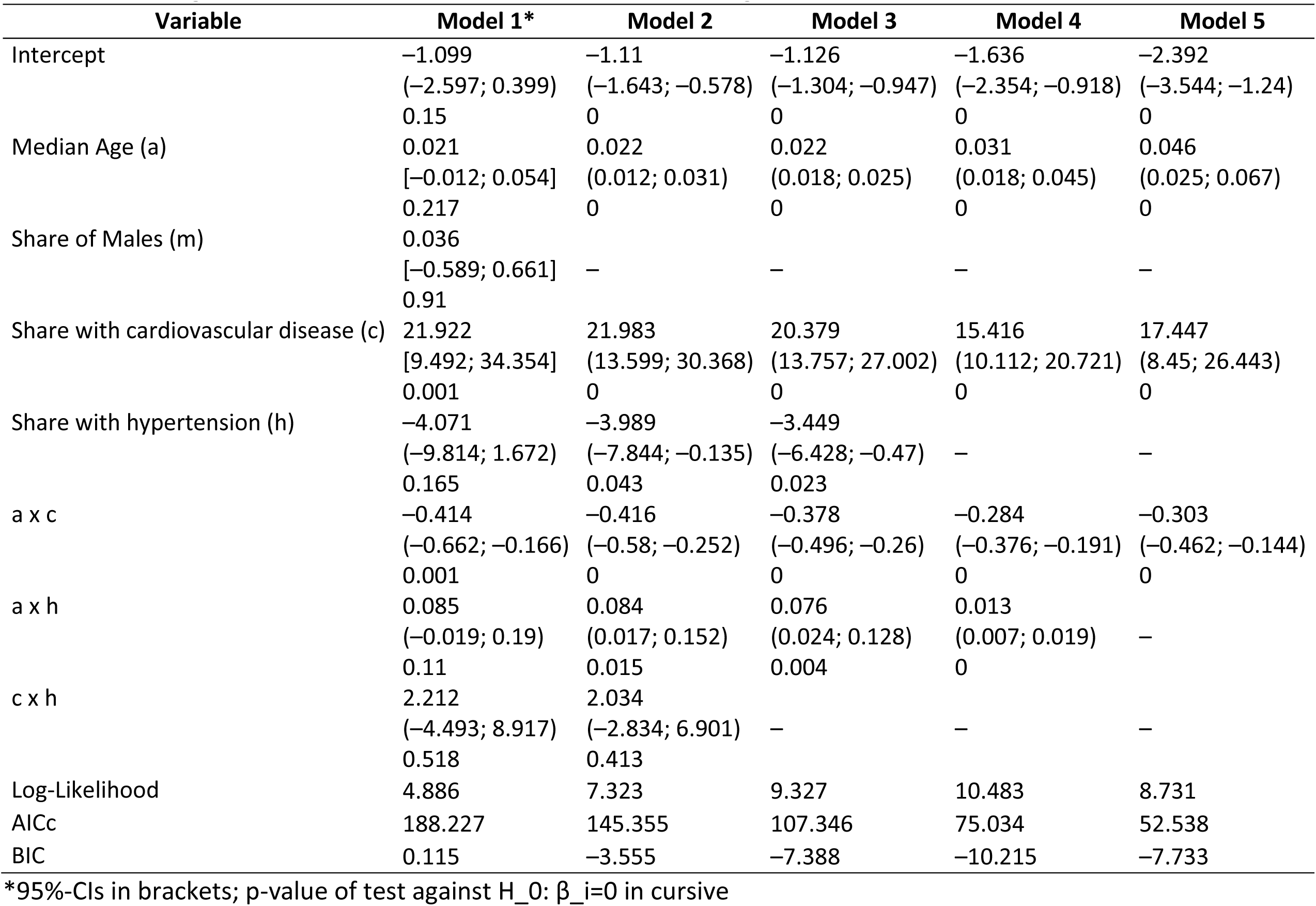
Meta-regression of associations between comorbidities, age, sex and COVID-death

### 3.4. Publicly available data

We found publicly available data that allowed to assess the outcomes presented above for both effects of comorbidities and age for Spain and France. Based on this data, the RR varies by age and sex (Table 3, A and B). Individuals aged 70+ years were at higher risk of death and hospitalisation than younger individuals (Table 3, A). For example, as of 21 May 2020 in Spain a 50-69 year-old male case of COVID-19 had an estimated risk of dying 6.9 (95% CI 6.0-8.0) times that of a male below 50 years of age. In terms of sex, men were at higher risk of dying or witnessing a severe course of the infection than women do (Table 3, B). For instance, in Italy a male aged 50 years or younger was estimated to have a 3.33 (95% CI 2.6-4.2) higher average risk of death after infection than a woman at the same age does.

**Table 3.**
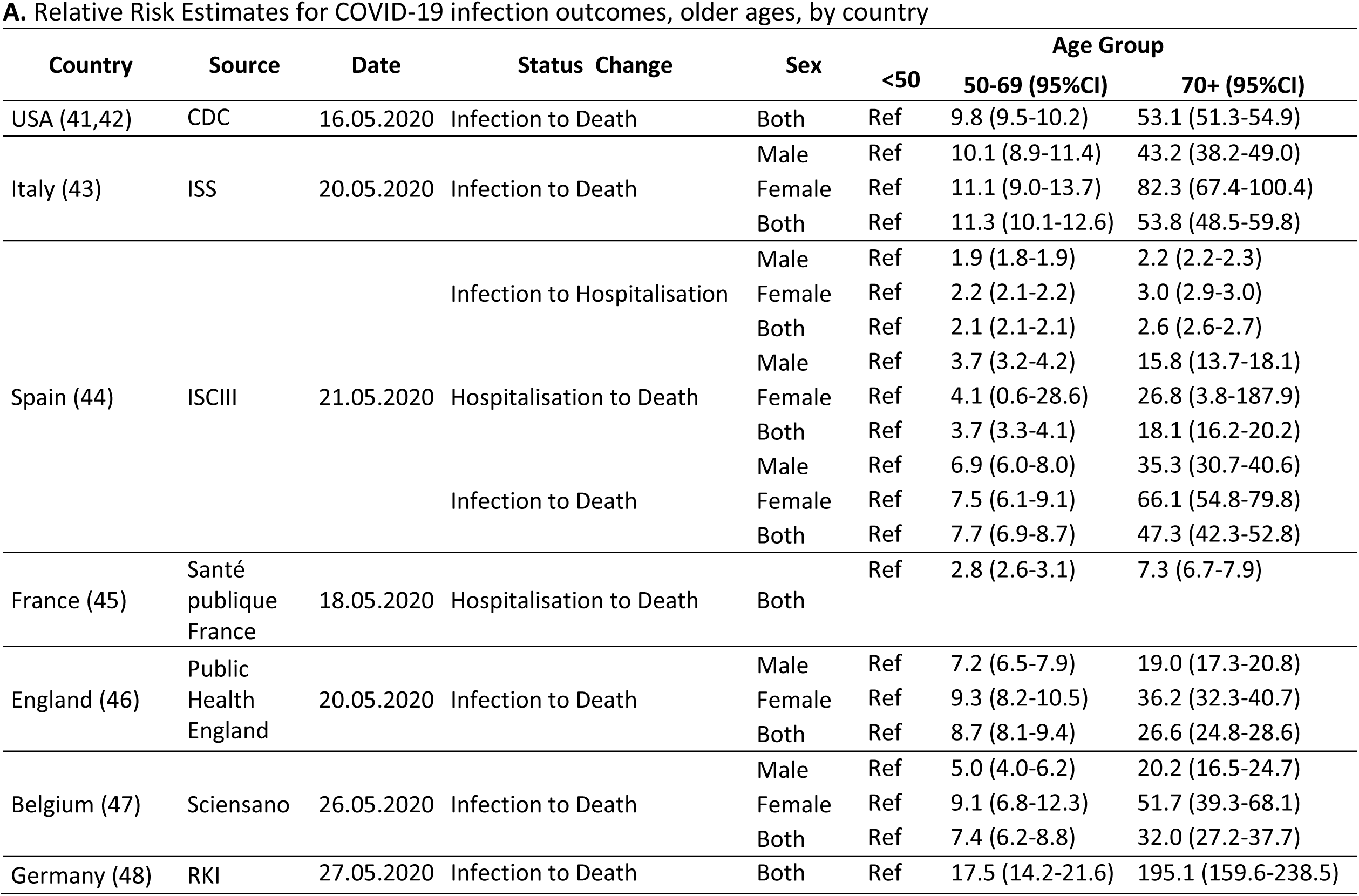
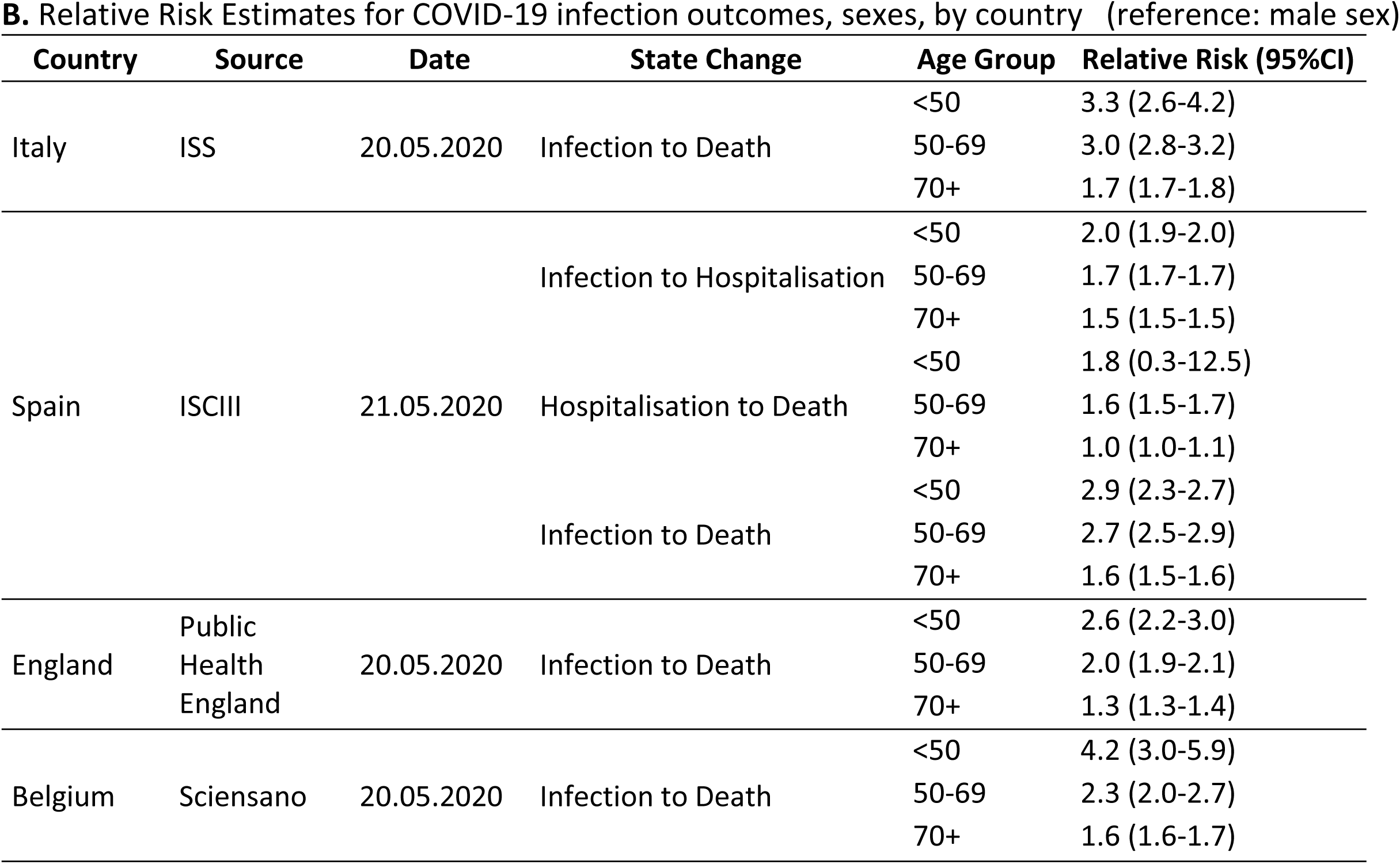
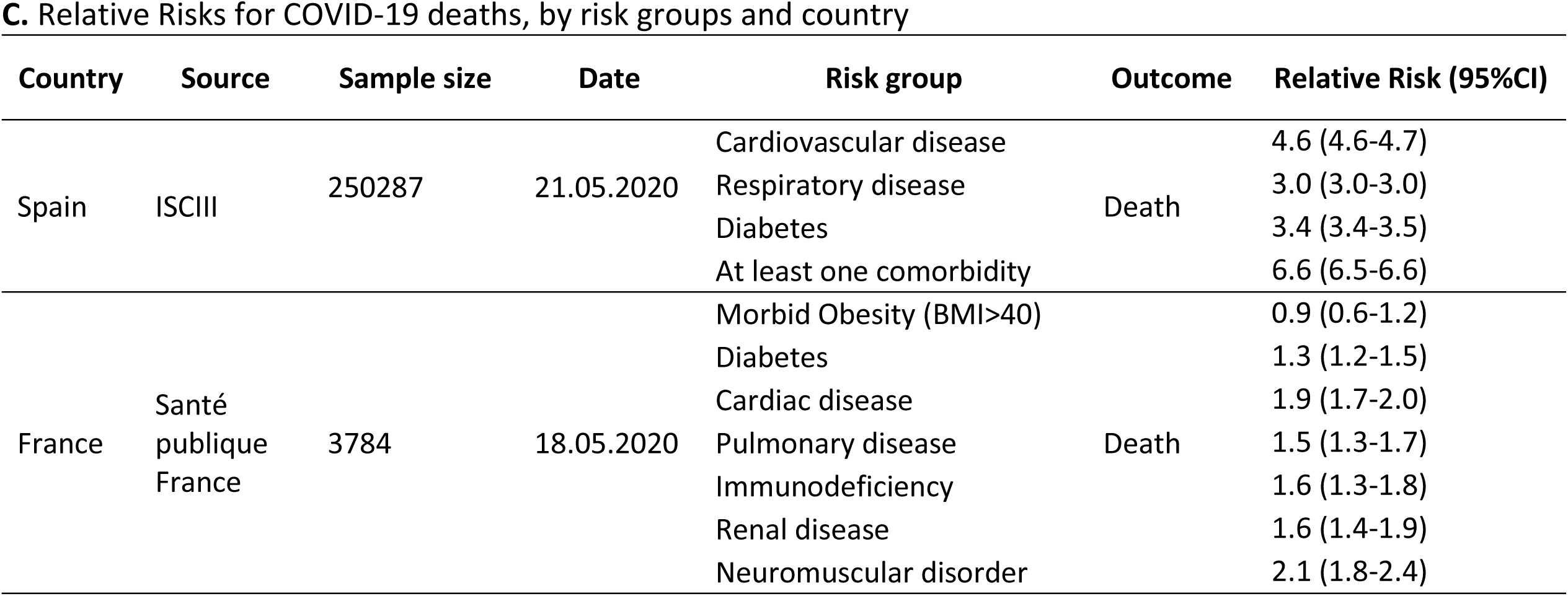
Relative Risk estimates based on publicly available data, reported from country health institutions, and others

Regarding comorbidities (Table 3, C), the RR of diabetes ranged between 1.3 (95% CI 1.2-1.5) and 3.4 (CI 3.4-3.5). In terms of cardiac and cardiovascular disease, the RR of death was 1.9 (CI 1.7-2.0)-4.6 (CI 4.6-4.7). For pulmonary and respiratory disease, the risk of death were 1.5 (CI 1.3-1.7) – 3.0 (CI 3.0-3.0).

## 4. Discussion

Results of our meta-analysis add to existing rather limited systematic reviews and meta-analyses (17, 19–22) by providing a more comprehensive analysis of the magnitude of risk associated with both comorbidities and further risk factors to hospitalisation, ICU admission, and death. We believe that the results have public health implications in four main fields.

First, we show that risk of dying from COVID-19 is associated with the most prevalent existing comorbidities, such as cerebrovascular [RR 2.6 (95% CI 1.7-4.1)] and cardiovascular [RR 3.3 (95% CI 2.3-4.5)] diseases, hypertension [RR 2.7 (95% CI 2.1-3.4)], COPD [RR 2.4 (95% CI 2.0-3.0)], and renal disease [RR 2.5 (95% CI 1.8-3.4)], with a relative risk of 2-3. Knowing the exact magnitude of risk is important for a prioritisation both of measures that aim to protect vulnerable population groups and prevention strategies. Explanations for the RR increase are in some instances related to the pathophysiology of the disease itself. For cardiovascular disease and hypertension, for example, a dysregulated innate immune response was found to influence severe COVID-19 infections (52). In addition to that, it has been documented that the angiotensin-converting enzyme 2 (ACE2) has a vital role in the cardiovascular and immune systems, and it is involved in the heart function and the development of hypertension and diabetes mellitus (53). Moreover, Zheng et al. considered that a disease severity in patients with cardiovascular disease can be associated with increased secretion of ACE2(54). There is evidence that hypertensive patients may experience a decreased expression of ACE2, and consequently, an elevation of the angiotensin II levels that generates a severe manifestation of the disease (55). The same protein and its poor regulation may also explain the link between COPD and smoking in terms of COVID severity and mortality (31, 22). Additionally, exacerbations in COPD cases are triggered by viral infections and environmental conditions (57). For diabetes, the increased RR is explainable by reduced pulmonary function and a thickening of the pulmonary basal lamina (1,58). Other than for comorbidities, factors like sex determine the COVID risk and women might be protected by hormonal factors (8).

Second, our analysis shows that association of these comorbidities with hospitalisation and ICU admission are generally less strong for these same comorbidities and other risk factors and death. This also corresponds to public data from Europe, where the proportion of infected people of older age or of other risk groups is relevantly higher in those who died compared to those who were hospitalised. In Spain, for people infected and older than 70 years old, regarding sex, the risk of dying was higher than the risk of hospitalisation [RR 2.6 (95% CI 2.6-2.7) vs. RR 47.3 (95% CI 42.3-52.8)]. In terms of comorbidities, while the risk of those with cardiovascular conditions to die from COVID-19 is relevantly raised (RR 3.3, 95% CI: 2.3-4.5), the risk to be hospitalised or admitted to ICU is only moderately increased (RR 2.1, 95% CI1.6-2.4). Other studies have also found that for patients with cerebrovascular disease and with diabetes the risk increase of ICU admission was lower than the risk of dying compared to other population groups (19,59). This finding is important in that implies that public health measures to protect healthcare surge capacities should not be equalled with measures to protect the vulnerable from death in the fight against COVID-19. Even if – hypothetically - fully protecting all people vulnerable from death, the effect on healthcare surge capacities of these same measure, measured by hospital beds, critical care beds, healthcare workers, and healthcare expenditure (60), will not be equally effective.

Third, we provide evidence for previously discussed risk factors of COVID-19 severity. Similarly to others (5,8), we observed low strength of association for obesity, and smoking. Interestingly, our meta-analyses showed less risk of severe outcomes of COVID-19 for healthcare professionals. This might be explained by a lower likelihood for underreporting in this population group but also by the healthy worker effect (61). Therefore, comparative studies on this occupational group are needed.

Fourth, we confirm the results of previous meta-regressions on the effect of age on COVID-19 outcomes and comorbidities (22). For several comorbidities (cardiovascular disease, hypertension, and diabetes) we showed weaker associations with deaths among study participants which were part of studies with a higher median age of patients. This implies that differentiation by specific predisposing conditions might be particularly effective in the lower age groups. This is also applicable as older patients suffer from multiple and coexisting medical conditions, reducing standalone effects of single conditions (22,62,63).

The limitations of our work derive from the restricted search, which was a rapid approach in one main data base of medical literature. With this review type, we sought for contextualised evidence to inform decision makers in terms of vulnerable population groups. Moreover, we did not include articles in Chinese language into our search due to lack of interpreters, and that could have affected our included studies and findings. The nature of the included data were often based on hospital recording implying bias in a sense that more severely symptomatic patients are more likely included. Although our assessment revealed high to moderate study quality, studies based on hospital records are highly selective regarding the population included. Regarding our analyses, we did not consider age groups separately due to wide ranges and inconsistencies in reporting from the studies, which would have implied major assumptions for our meta-analyses. However, we approached this by assessing effect modification of age on different comorbidities and found effect modification for several comorbidities. The meta-analyses we conducted is univariate, and we did not adjust for co-morbidities that appear in parallel.

Generalisability and country-comparisons of results from publicly available data are limited by different data sources used. Also, our effect estimates for some countries only investigate subgroup of hospitalised cases (e.g. France), which may lead to a systematic underestimation of the effect estimates, since these individuals likely have an increased risk of severity. However the qualitative implications of our estimates seem to be robust across different geographic regions as well as different data sources.

In conclusion, we provide evidence synthesis for the exact magnitudes of effects of comorbidities on severity of disease in COVID-19 to help target public health measures more towards individual population groups at risk. Most importantly, we show that the mortality risk from COVID-19 is associated with the most prevalent existing comorbidities, such as cerebrovascular and cardiovascular diseases, hypertension, and renal disease and that there is a decrease of the effect of comorbidities on severity of disease with increasing age for some comorbidities, and that there is a generally higher strength of association in these comorbidities for death than for other severe course of disease.

## Data Availability

Available publications on COVID-19 and other publicly available sources were employed in this manuscript.

## Footnotes

1 US, Italy, Spain, France, England, Belgium, and Germany.

2 ≈ *β*_*axh*_ * *a* * Δ*h* ≈ 0.013 * 50.0 * 01

3 ≈ 0.013 * 70 * 0.01

